# Molecular analysis of several in-house rRT-PCR protocols for SARS-CoV-2 detection in the context of genetic variability of the virus in Colombia

**DOI:** 10.1101/2020.05.22.20107292

**Authors:** Diego A. Álvarez-Díaz, Carlos Franco-Muñoz, Katherine Laiton-Donato, José A. Usme-Ciro, Nicolás D. Franco-Sierra, Astrid C. Flórez, Sergio Gómez Rangel, Luz Dary Rodríguez, Juliana Barbosa, Erika Ospitia, Diana Marcela Walteros, Martha Lucia Ospina Martínez, Marcela Mercado-Reyes

## Abstract

The COVID-19 pandemic caused by SARS-CoV-2 is a public health problem unprecedented in the recent history of humanity. Different in-house real-time RT-PCR (rRT-PCR) methods for SARS-CoV-2 diagnosis and the appearance of genomes with mutations in primer regions have been reported. Hence, whole-genome data from locally-circulating SARS-CoV-2 strains contribute to the knowledge of its global variability and the development and fine tuning of diagnostic protocols. To describe the genetic variability of Colombian SARS-CoV-2 genomes in hybridization regions of oligonucleotides of the main inhouse methods for SARS-CoV-2 detection, RNA samples with confirmed SARS-CoV-2 molecular diagnosis were processed through next-generation sequencing. Primers/probes sequences from 13 target regions for SARS-CoV-2 detection suggested by 7 institutions and consolidated by WHO during the early stage of the pandemic were aligned with Muscle tool to assess the genetic variability potentially affecting their performance. Finally, the corresponding codon positions at the 3′ end of each primer, the open reading frame inspection was identified for each gene/protein product. Complete SARS-CoV-2 genomes were obtained from 30 COVID-19 cases, representative of the current epidemiology in the country. Mismatches between at least one Colombian sequence and five oligonucleotides targeting the RdRP and N genes were observed. The 3’ end of 4 primers aligned to the third codon position, showed high risk of nucleotide substitution and potential mismatches at this critical position. Genetic variability was detected in Colombian SARS-CoV-2 sequences in some of the primer/probe regions for in-house rRT-PCR diagnostic tests available at WHO COVID-19 technical guidelines; its impact on the performance and rates of false-negative results should be experimentally evaluated. The genomic surveillance of SARS-CoV-2 is highly recommended for the early identification of mutations in critical regions and to issue recommendations on specific diagnostic tests to ensure the coverage of locally-circulating genetic variants.

**HIGHLIGHTS:**

- Colombian SARS-CoV-2 sequences displayed genetic variability in some target regions used for COVID-19 diagnosis.
- Mismatches in critical primer regions could impact their performance and the rate of false negative results.

## INTRODUCTION

In late December 2019 in Wuhan city (China), a new coronavirus called SARS-CoV-2 (initially nCoV-2019) caused the outbreak of a respiratory disease known as the infectious disease due to the new coronavirus (COVID-19) (Zhou et al., 2020; Zhu et al., 2020). Soon after learning about the potential for transmission of this virus in the context of a globalized world, countries took swift and extreme measures such as border closings, rigorous follow-up of contacts, and mandatory preventive isolation. In Colombia, the first case of COVID-19 was identified on March 6, 2020, shortly before the World Health Organization (WHO) declared COVID-19 as a pandemic after it had spread in 114 countries in all continents and having claimed the lives of 4,291 people (WHO, 2020a).

The first case of COVID-19 in Colombia was imported from Italy, a country that had the most alarming epidemic peak in Europe at that time. Shortly afterwards, cases of COVID-19 were diagnosed in travelers from other origins, as well as in multiple of their contacts. On April 20, 2020, community transmission cases already exceeded 10% of the total cases registered in the country, which is why the transition to the mitigation phase was declared. Until May 18, 2020, 4,629,000 cases and 297,380 deaths have been reported globally, and in Colombia, 16,295 cases and 592 deaths have been reported (Dong, Du, & Gardner, 2020).

SARS-CoV-2 is a betacoronavirus with a positive polarity single-stranded RNA genome of approximately 30 kb. This new coronavirus shares a global identity of 96.2% with the bat coronavirus RaTG13 (Zhou et al., 2020), 91.2% with the Malay Pangolin coronavirus isolate, Pangolin-CoV (Zhang, Wu, & Zhang, 2020), and even 97.5% with RmYN02 derived from bat when the ORF1ab gene is exclusively analyzed (Wang, Pipes, & Nielsen, 2020). Its recent origin is enigmatic since there is a high similarity between the amino acid sequence in the receptor-binding domain (RBD domain) of subunit 1 (S1) of the Spike protein of SARS-CoV-2 and that of Pangolin-CoV, but the latter lacks the polybasic furin processing site, exclusive to SARS-CoV-2 (Andersen, Rambaut, Lipkin, Holmes, & Garry, 2020). Accumulated evidence suggests a zoonotic origin of the virus as a result of recombination with a yet unidentified coronavirus or convergent evolution driven by natural selection to optimize interaction with the human ACE2 cell receptor (Wang et al., 2020; WHO, 2020b). After the publication of the complete SARS-CoV-2 genome in genomic data repositories such as NCBI (MN908947.3) and GISAID in mid-January 2020, health agencies and researchers from different countries quickly developed SARS-CoV-2 screening tests based on realtime RT-PCR (rRT-PCR) that amplify different SARS-CoV-2 gene regions. Hundreds of commercial kits are under development and many of them have been licensed for emergency use (https://www.finddx.org/covid-19/pipeline/). Various in-house protocols were also developed and published on the website of the World Health Organization (WHO) for informational purposes without implying endorsement, preference or validation by this entity (WHO, 2020c). However, most of these protocols were published during January 2020 when only 230 virus sequences were available that circulated exclusively in Asia and Europe, except for a few cases in the United States and Canada (Holshue et al., 2020).

Since then, refinements of these protocols are not known in the context of nearly 30,000 sequences reported on May 18, 2020, worldwide, including Latin America, which provide a more complete perspective of the accumulated genetic variability and sequence particularities of viruses circulating in specific regions that could affect the efficiency and sensitivity of the rRT-PCR protocols currently shared by WHO. The mutation rate of SARS-CoV-2 as a virus with an RNA genome is higher than that of viruses with a DNA genome (Tang et al., 2020), with an estimated mean evolutionary rate of 2.24 x 10^-3^ substitutions/site/year (Li, Li, Cui, & Wu, 2020); therefore, changes in the sequence could occur over time that compromise the operational performance of diagnostic tests (PAHO, 2020). The objective of this study was to describe the genetic variability of Colombian SARS-CoV-2 genomes in hybridization regions of oligonucleotides of the main in-house methods for SARS-CoV-2 detection.

## MATERIALS AND METHODS

### Patients and samples

Nasopharyngeal swab samples from patients with suspected SARS-CoV-2 infection were received at the Instituto Nacional de Salud (INS) as part of the virological surveillance of COVID-19 from 11 Colombian departments and the capital district. According to the national law 9/1979, decrees 786/1990 and 2323/2006, the INS is the reference lab and health authority of the national network of laboratories and in cases of public health emergency or those in which scientific research for public health purposes as required, the INS may use the biological material for research purposes, without informed consent, which includes the anonymous disclosure of results. This study was performed in accordance with the ethical standards noted in the 1964 Declaration of Helsinki and its later amendments. The information used for this study comes from secondary sources of data that were previously anonymized and do not represent a risk to the community.

### RNA extraction and real-time rRT-PCR

Viral RNA was obtained using the automated MagNA Pure LC nucleic acid extraction system (Roche Diagnostics GmbH, Mannheim, Germany) and viral RNA detection was performed by rRT-PCR using the Superscript III Platinum One-Step Quantitative RT-kit. PCR (Thermo Fisher Scientific, Waltham, MA, USA), following the Charité-Berlin protocol (Victor M. Corman et al., 2020) for the amplification of the SARS-CoV-2 E and RdRp genes.

### Next generation sequencing

The complete SARS-CoV-2 genome sequence of 30 patients was obtained through NGS, ten genomes with Oxford Nanopore (Oxford Nanopore Technologies, Oxford, UK) and 20 genomes with lllumina MiSeq (lllumina, San Diego, CA, USA) technologies, following the artic.network “nCoV-2019 sequencing protocol” (Quick, 2020). In both strategies, SARS-CoV-2 specific oligonucleotides were used for the generation of amplicons by means of a Q5^®^ high fidelity DNA polymerase (New England Biolabs Inc., UK), in order to avoid the introduction of artificial mutations. The genomes were assembled by mapping to the reference genome (NC_045512.2) using the BWA (Li et al., 2020) and BBmap (brian-jgi, 2020) software to generate a consensus genome by the two assembly tools.

### Genetic diversity analysis

The Colombian genomes and oligonucleotides from of the in-house protocols were aligned with the Muscle tool (Edgar, 2004) using the MEGA X software (Kumar, Stecher, Li, Knyaz, & Tamura, 2018). Substitutions matrices of the Colombian genomes respect to the reference genome (NC_045512) at the nucleotide and amino acid levels were generated for the 13 rRT-PCR protocols published at the WHO website (WHO, 2020c), which several countries have established as their preferred diagnostic protocol for SARS-CoV-2.

### Oligonucleotides analysis

Thermodynamic features (priming Tm and ΔG at the variable sites, mispriming and ΔG, hairpin ΔG, and primer dimer ΔG) and the codon position at the 3’ end for oligonucleotides with conflicting sites and optimized oligonucleotides were evaluated into the PrimerSelect module of the LaserGene v8.1 suite (DNASTAR Inc. Madison, WI, USA).

## RESULTS

### Several target regions for SARS-CoV-2 molecular detection using in-house protocols

A total of 39 primer and probe sequences from the main in-house rRT-PCR protocols for SARS-CoV-2 detection published at WHO website were aligned to the reference sequence derived from the first confirmed case at Wuhan, Hubei province, China, and named Wuhan-1 strain (GenBank Accession Number: NC_045512.2). The protocols targeted 13 different genome regions inside the Orf1ab (Nsp9, NsplO, Nsp11, RdRp, ExoN), E (Envelope) and N (Nucleocapsid) genes with 61,5% (8/13) of the assays targeting the N gene (Figure 1).

**Figure 1.**
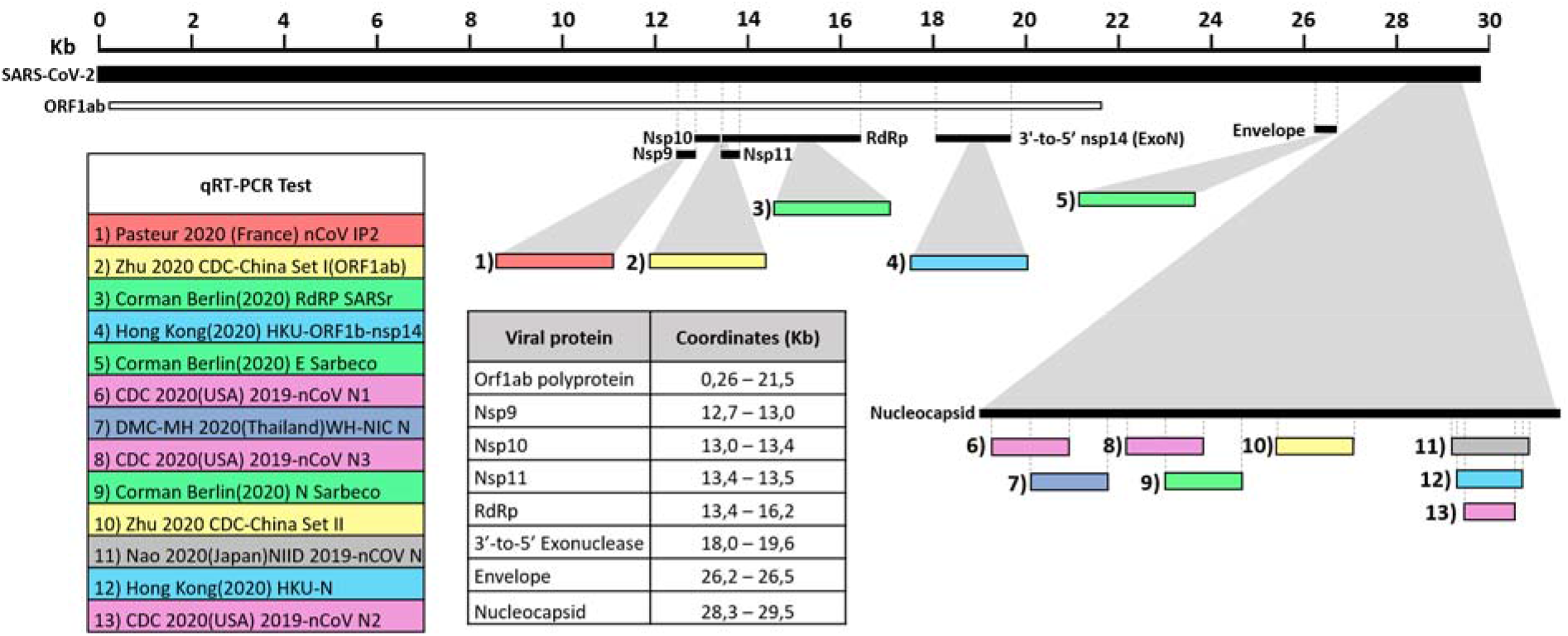
Target hybridization regions of the primers/probes employed by the principal in-house protocols for molecular detection of SARS-CoV-2. The protocols targeted 13 different genome regions inside the Orf1ab (Nsp9, Nsp10, Nsp11, RdRp, ExoN), E (Envelope) y N (Nucleocapsid) genes. Target hybridization regions of the primers / probes employed by the principal in-house protocols for molecular detection of SARS-CoV-2. The different genes and protein products, as well as the coordinates in kilobases (Kb) of the genes and protein products to which they are directed were estimated according to the SARS-CoV-2 reference genome available at GenBank (NC_045512.2).

### Point mutations in Colombian SARS-CoV-2 genomes at the target hybridization sequences of some inhouse protocols for SARS-CoV-2 detection

A total 30 whole genome SARS-CoV-2 sequences from Colombia were included in the previously obtained alignment for genetic variability analysis. These sequences derived from cases with confirmation dates from March 6^th^-24^th^. 2020 were remitted from 11 departments and the capital district (Antioquia, Bogotá D.C., Bolivar, Caldas, Cauca, Magdalena, Norte de Santander, Quindío, Risaralda, Santander, Tolima and Valle del Cauca) through the National Public Health Laboratories Network to the Colombian National Institute of Health for diagnostic confirmation.

From the alignment of the 39 oligonucleotides to the reference and Colombian SARS-CoV-2 sequences, 5 showed mismatches with at least one Colombian sequence (Table 1). The conflicting sites in the primer/probe sequences were due to 1) a mismatch between the oligonucleotide and the reference and Colombian sequences or 2) a mismatch between the oligonucleotide and one or more Colombian sequences.

**Table 1.**
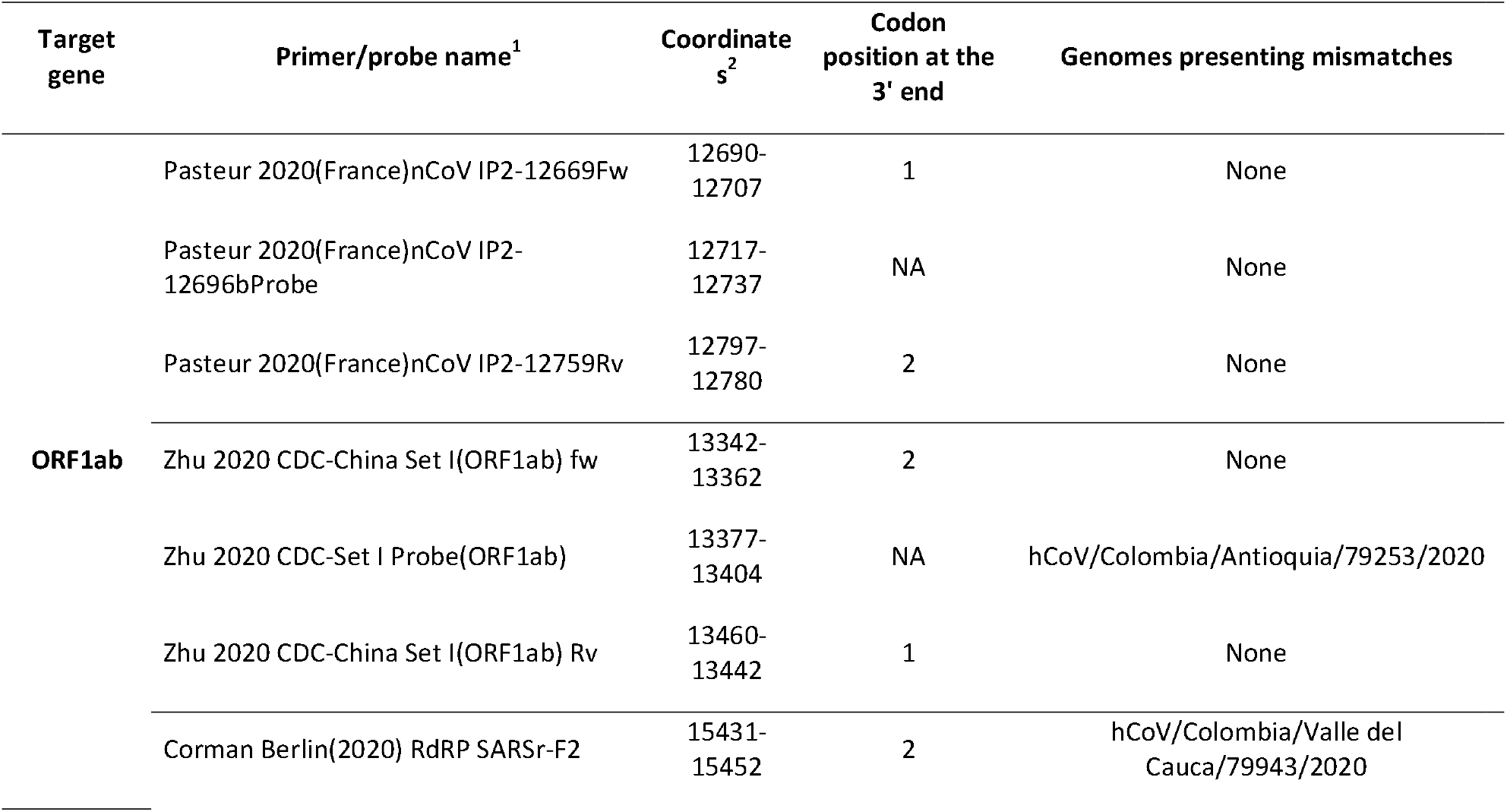
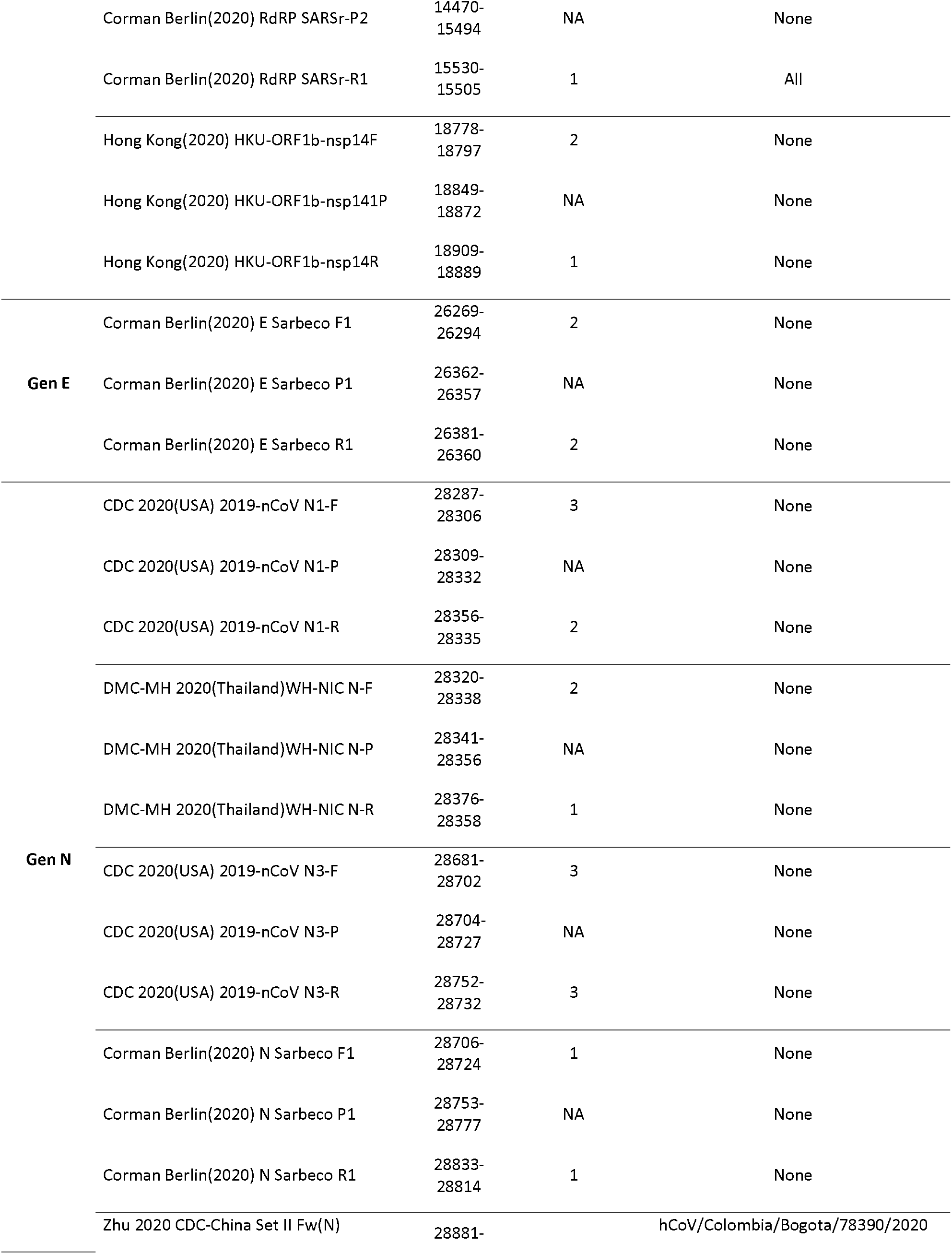
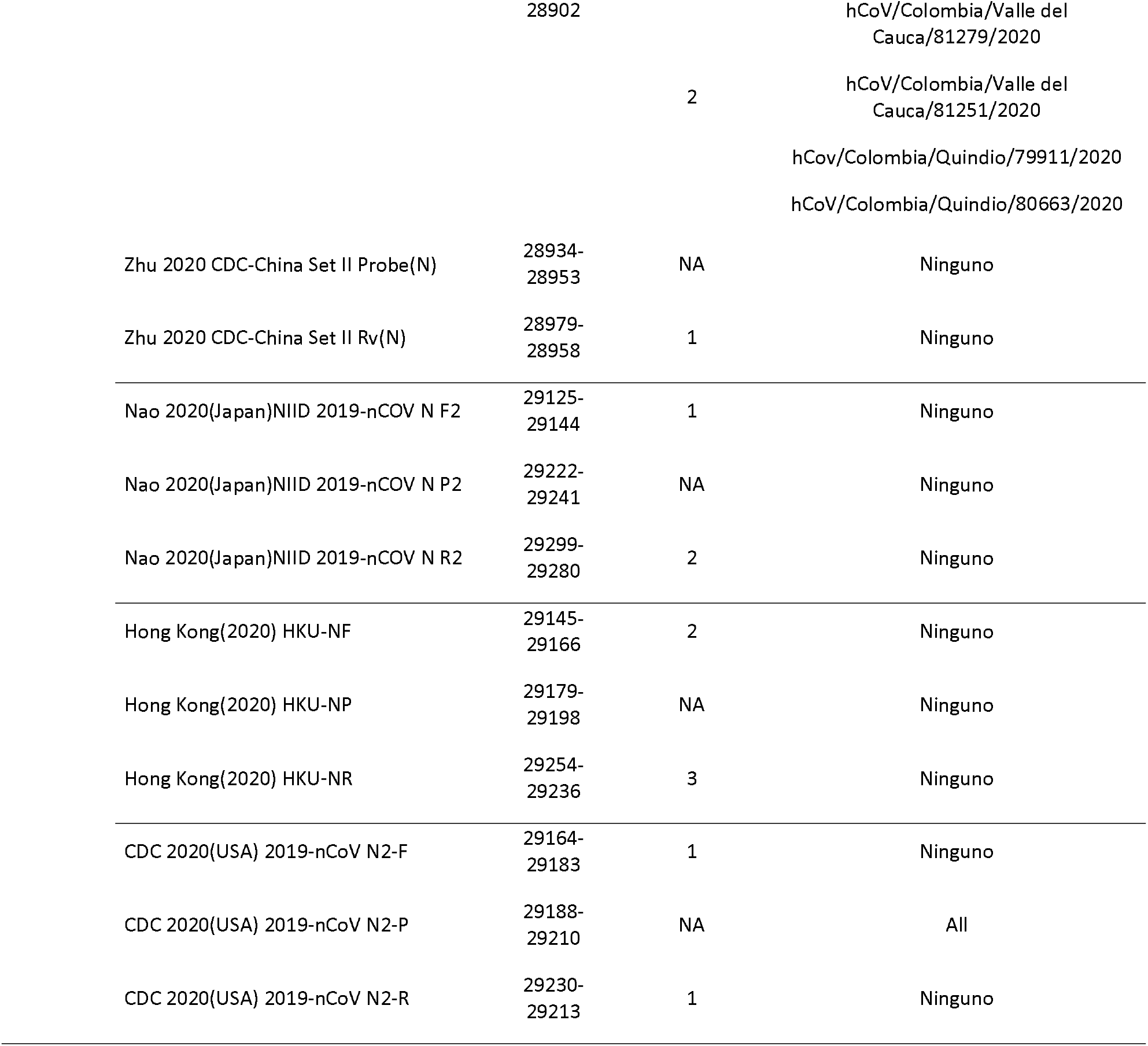
Target gene/regions and design conflicts of the analyzed primers and probes according to the genetic diversity of the Colombian SARS-CoV-2 strains.

The Hong Kong(2020) HKU-NP probe (Chu et al., 2020) (Figure 2) was extremely different to all the reference and Colombian SARS-CoV-2 by four nucleotide sites. This probe also presents the formation of a highly stable hairpin and self-dimer structures (Table S1). The Corman Berlin(2020) RdRP SARSr-F2 (Victor M. Corman et al., 2020) (Figure 3) region was variable in one Colombian SARS-CoV-2 sequence from the department of Valle del Cauca (ID: 79943), involving a critical site at the 3’ pentamer. This site is supposed to prevent the correct hybridization of the 3’ end of the primer, leading to inefficient or unsuccessful extension by the DNA polymerase. The RdRP SARSr-R1 primer (Victor M. Corman et al., 2020) showed a degenerate site which does not comprise the nucleotide found in the reference and Colombian SARS-CoV-2 sequences. However, this mismatch was located at an internal site of the primer, only partially affecting the thermodynamic profile of the primer-target hybridization. The Zhu 2020 CDC-China Set I Probe(ORF1ab) (Zhu et al., 2020) (Figure 4) was found to be almost completely complementary with the Colombian SARS-CoV-2 sequences, except for a viral sequence obtained from a human case in the department of Antioquia (ID: 79253), bearing a single substitution at the seventh probe position, without considerable effect on the thermodynamic features for probe-target hybridization (Table S1). The Zhu 2020 CDC-China Set II Fw(N) primer (Figure 5) hybridization was found to be critically affected by the accumulated genetic diversity of the Colombian SARS-CoV-2 strains. At the 5’ end of the primer a triple-nucleotide substitution GGG→AAC in three sequences from Bogotá and Valle del Cauca affected the Tm and ΔG. At the 3’ region, two sequences from Quindío displayed a substitution affecting the 3’ pentamer stability.

**Figure 2.**
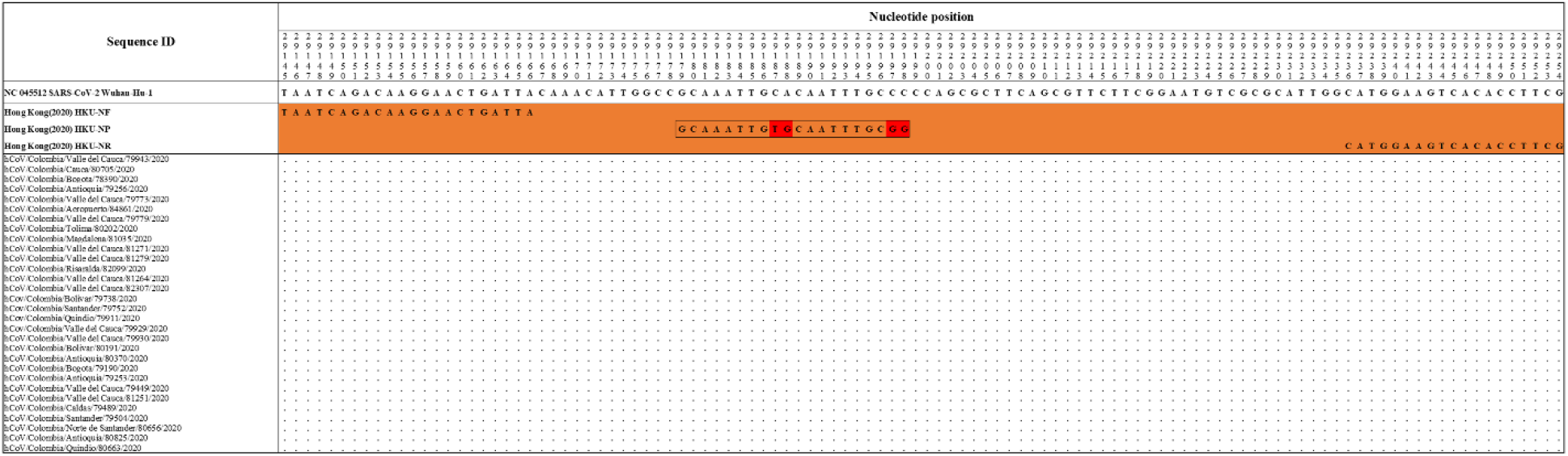
Genetic diversity at the target region of the Hong_Kong (2020)_HKU-NP assay. Hong Kong (2020) _HKU-NP probe displayed mismatches at positions 29187-88 and 29197-98 (highlighted in red), with all the Colombian sequences and the RefSeq displaying CA and CC, respectively. Genomic positions were estimated according to the SARS-CoV-2 reference genome available at GenBank (NC_045512.2).

**Figure 3.**
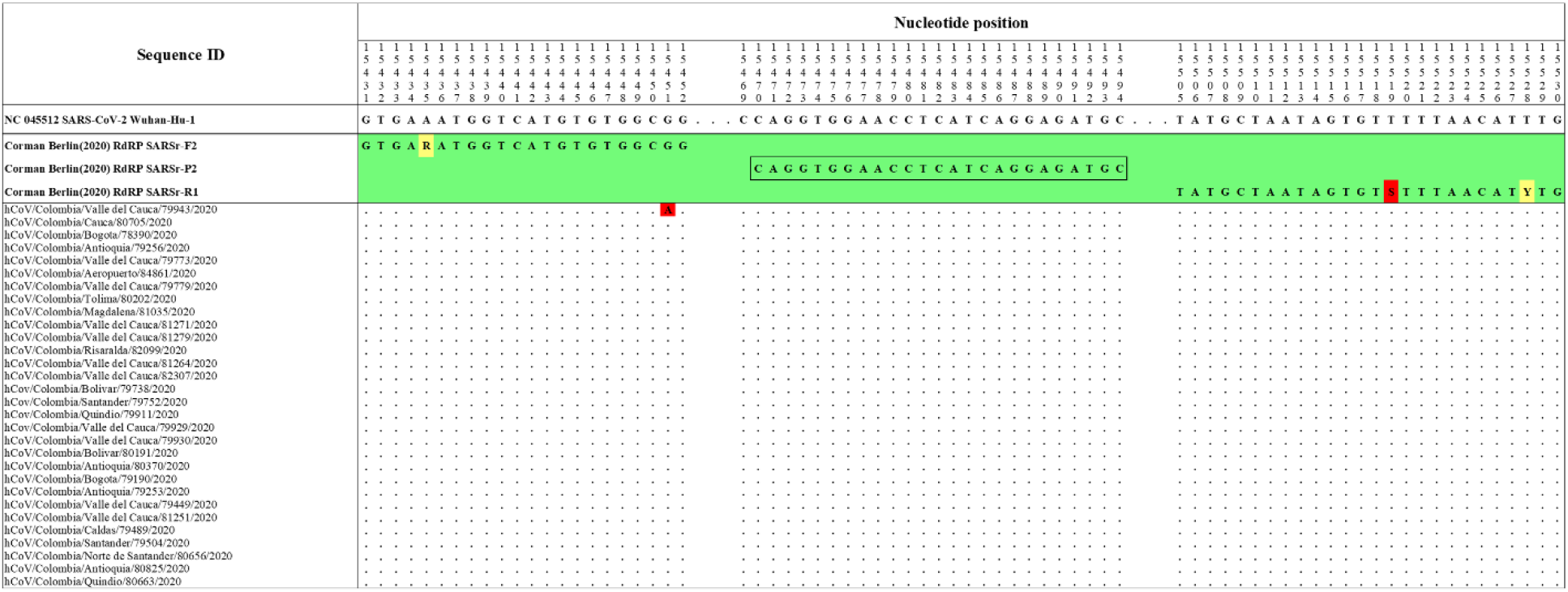
Genetic diversity at the target region of the Corman_Berlin (2020) _RdRP assay. The Colombian genome hCoV/Colombia/Valle_del_Cauca/79943/2020 displayed a substitution (G to A) at position 15,451 (highlighted in red), affecting the primer Corman_Berlin (2020) _RdRP_SARSr-F2 hybridization region. The substitution is located in the penultimate position at the 3 ‘end of the promer, generating a mismatch that could interfere with the 3’ pentamer stability. A degenerate base (S = C or G) in the primer Corman_Berlin (2020) _RdRP_SARSr-R1 at position 15,519 (highlighted in red) produces a mismatch with all the Colombian genomes and the RefSeq as they have T in the sense sequence, so the degenerate base should be one that includes an A among degenerate alternatives. Genomic positions were estimated according to the SARS-CoV-2 reference genome available at GenBank (NC_045512.2).

**Figure 4.**
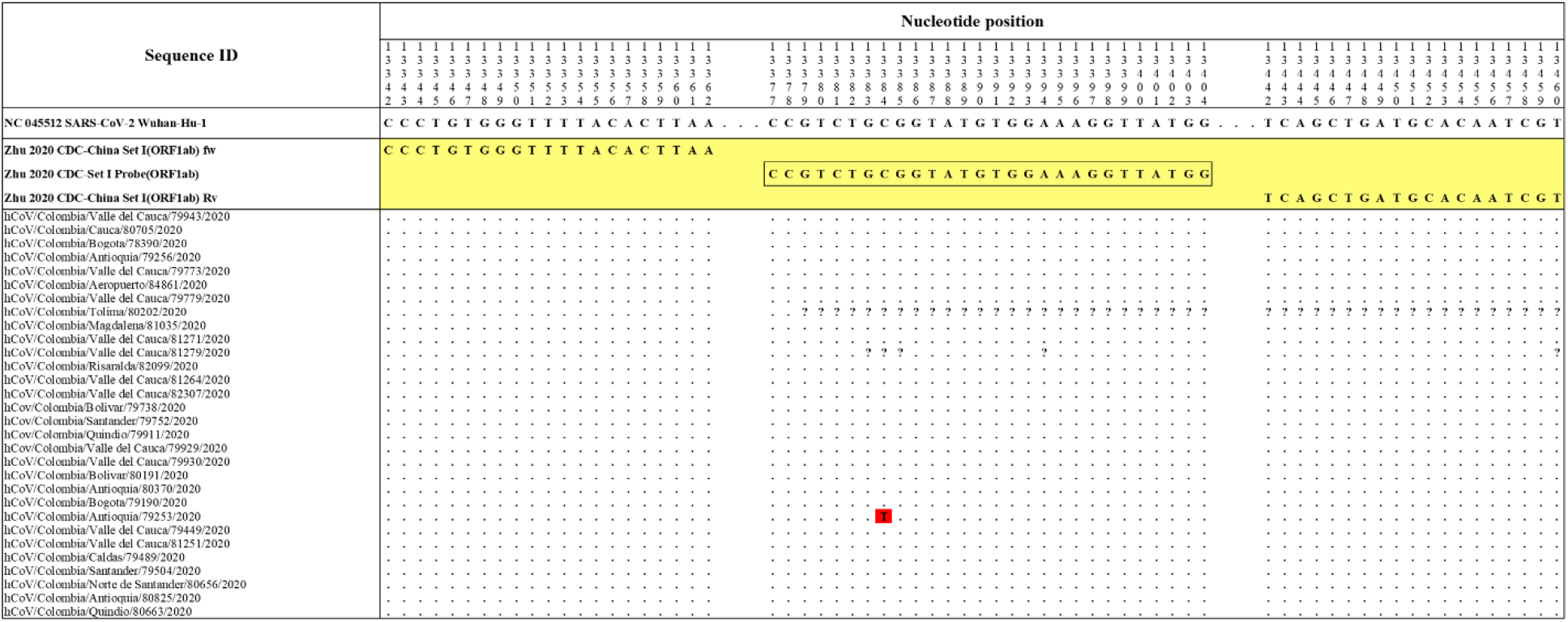
Genetic diversity at the target region of the Zhu_2020_CDC-Set I assay. The Colombian genome hCoV/Colombia/Antioquia/79253/2020 displayed a substitution (C to T) at position 13,384 (highlighted in red) where the probe Zhu_2020_CDC-Set I Probe (ORFlab) hybridizes. Genomic positions were estimated according to the SARS-CoV-2 reference genome available at GenBank (NC_045512.2).

**Figure 5.**
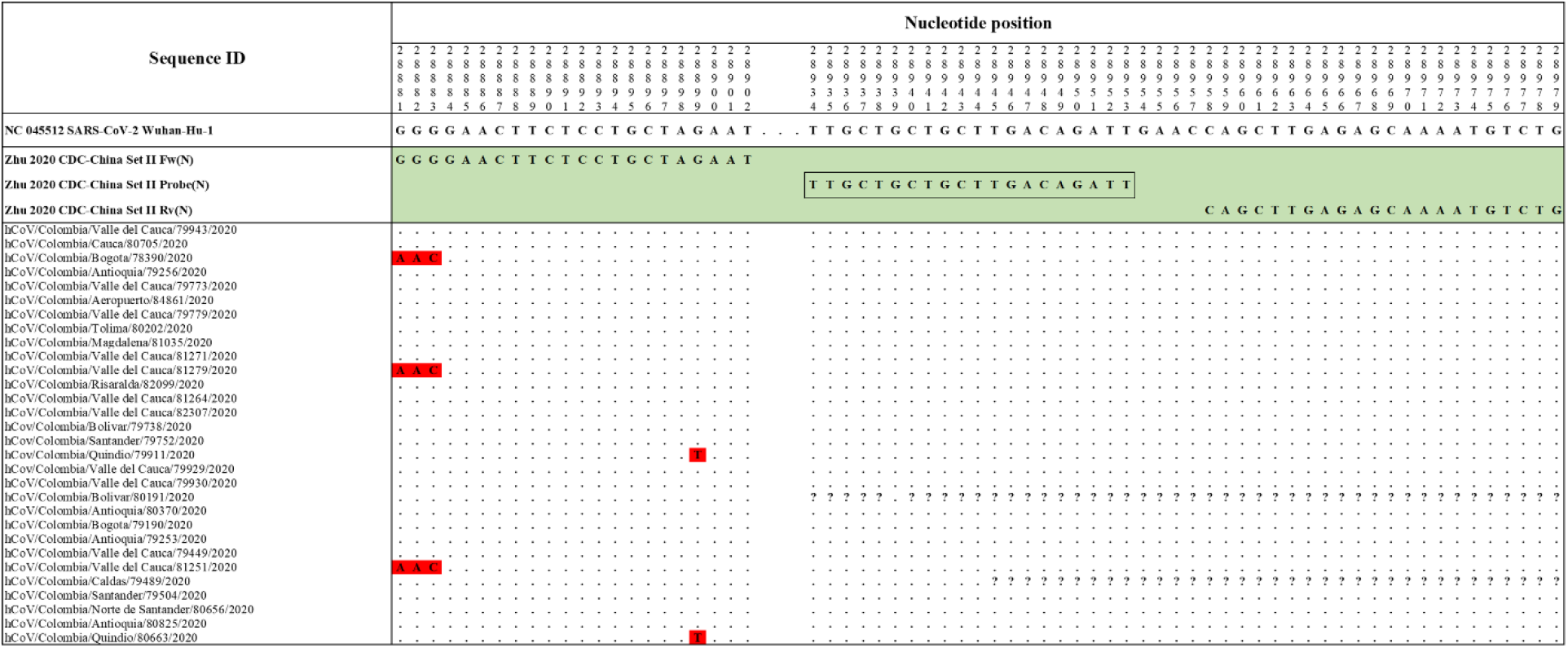
Genetic diversity at the target region of the Zhu_2020_CDC-China_Set II assay. Three Colombian genomes, hCoV/Colombia/Bogota/78390/2020, hCoV/Colombia/Valle_del_Cauca/81279/2020 and hCoV/Colombia/Valle_del_Cauca/81251/2020 have the same pattern of substitutions (GGG to AAC) in primer Zhu_2020_CDC-China_Set II Fw (N) in the first three nucleotides at the 5’ end (highlighted in red). Furthermore, the hCoV/Colombia/Quindio/79911/2020 and hCoV/Colombia/Quindio/80663/2020 genomes displayed a substitution (G to T) at position 28,899 where this oligonucleotide hybridizes (highlighted in red). Genomic positions were estimated according to the SARS-CoV-2 reference genome available at GenBank (NC_045512.2).

The primers and probes sets proposed by Pasteur 2020 (France)nCoV IP2 (Fig. S1), Hong Kong (2020) HKU-ORFlb-nspl4 (Fig. S2), Corman Berlin(2020) E Sarbeco (Fig. S3), CDC 2020(USA) 2019-nCoV N1 (Fig. S4), DMC-MH 2020(Thailand)WH-NIC N (Fig. S4), CDC 2020(USA) 2019-nCoV N3 (Fig. S5), Corman Berlin(2020) N Sarbeco (Fig. S5), Nao 2020(Japan)NIID 2019-nCOV N (Suppl. Fig. 6) and CDC 2020(USA) 2019-nCoV N2 (Fig. S6) showed correspondence with the reference sequence and with all the accumulated genetic variability in available sequences of Colombian strains of SARS-CoV-2.

### The third codon position was found to align with the 3’ end of some primers with intended use for molecular detection of SARS-CoV-2

Codon positions in coding regions are differentially susceptible to nucleotide substitution, being the third codon position associated with a higher substitution rate. On the other hand, the perfect matching of the last nucleotide at the 3’ end of every forward and reverse primer is critical for DNA polymerase-based extension during PCR amplification (Staheli, Ryan, Bruce, Boyce, & Rose, 2009). Therefore, the rational design of primer to be used with rapidly evolving RNA viruses should have the requisite of avoiding third and sometimes first codon positions.

Codon position of the last nucleotide at the 3’ end of every analyzed primer was identified according to the corresponding open-reading frame. The 3’ end of 11 primers corresponded to the first codon position, another 11 primers had 3’ ends located at the second codon position and the 3’ end of 4 primers aligned with the third codon position (Table 1). One of the primers (Nao 2020(Japan)NIID 2019-nCOV N F2) had the 3’ end aligned with the first codon position of the codon CUA which encodes for alanine. This codon allows a substitution at the first codon position (C to U) to be synonymous and therefore may escape to the selection pressure at the protein level.

The 3’ end of the primers CDC 2020(USA) 2019-nCoV N1-F, CDC 2020(USA) 2019-nCoV N3-F, CDC 2020(USA) 2019-nCoV N3-R and Hong Kong(2020) HKU-NR aligned with the third codon position of their corresponding ORFs. Two of these primers were the forward and reverse primers proposed to amplify a specific target (CDC 2020(USA) 2019-nCoV N3) at the N gene making this protocol very susceptible to false negative results as the viral genetic variability increases.

## Discussion

Next Generation Sequencing (NGS) technologies are a valuable tool in determining the whole genome of microorganisms impacting on public health, they have accelerated our ability to understand important factors in the biology of infectious diseases such as the identification of determinants of virulence, drug-resistance associated substitutions, vaccine design and genomic epidemiology, a powerful approach to integrate epidemiologic and genetic information in reconstructing transmission patterns and infectious disease dynamics (Gwinn, MacCannell, & Armstrong, 2019). NGS has also enabled pathogen discovery in a timely manner compared to traditional methods, being essential to the virus taxonomic classification during the beginnings of COVID-19 pandemics (Zhou et al., 2020; Zhu et al., 2020). This robust molecular information is also the raw material for the development and refinement of molecular and serological diagnostic methods (Wang et al., 2020) and all sequence-based designs are susceptible to improvement as the virus disseminates and its genetic variability increases.

Mutation is the fundamental source of genetic variation and RNA viruses are particularly susceptible to have high mutation rates during the genome replication (Sanjuan, Nebot, Chirico, Mansky, & Belshaw, 2010). Therefore, RNA viruses display high substitution rates when analyzed through time. Although sequence identity between the analyzed genomes and the Wuhan reference strain (NC_045512) was around 99.9%, the estimate substitution rate for SARS-CoV-2 is in the range of 1.67-4.67 x 10^-3^/site/year (Tang et al., 2020). Therefore, it is evident that the genomic data of this study allow the refinement or even the design of more precise and efficient protocols for the molecular detection of the genetic variants of SARS-CoV-2 circulating in Colombia.

An enormous effort of every official and private laboratory around the word has led to the availability of hundreds of commercial kits and in-house protocols for the molecular detection and COVID-19 diagnostic confirmation. While this effort is welcome and some recommendations have been provided by regional agencies, the availability of all these methods imply the need for an evidence-based criterion for decision-making about the best option at country level. In this study, based on NGS data of complete SARS-CoV-2 genomes from Colombian cases, timely evidence is provided on the genetic variability of representative strains of the circulating viruses in the country that should be considered for an evidence-based improvement of the routine diagnostic tests.

The majority of the evaluated in-house molecular assays displayed genetic stability at the primers/probes target regions. However, some these oligonucleotides displayed mismatches that were considered of minor or major importance for the test performance based on the in silico analysis. Some of the analyzed primers/probes displayed mismatches when compared to target sequences of some Colombian SARS-CoV-2 strains. Polymorphic sites in the complete SARS-CoV-2 genomes obtained in this study, were supported by sequencing depths between 123 and 390X sufficient to identify these sequence variations unequivocally. The analyzed genome sequences corresponded to the early introduction and dispersion of the virus in the country. It is expected that some of the identified nucleotide substitutions remain stable in subsequent transmission chains of the corresponding clusters of cases, as demonstrated in the present study as character states shared by two or more sequences.

Prior to the arrival of SARS-CoV-2 in Colombia, the Charité-Berlin protocol described by Corman and collaborators was established as a routine in detection (Victor M. Corman et al., 2020) following the recommendation of the Pan American Health Organization (PAHO). Their three molecular targets (RdRp, E and N genes) were analyzed and RdRp displayed some conflicting nucleotide sites. The antisense primer RdRp SARSr R1 presented a degenerate site at the nucleotide position 15,519 equivalent to nucleotides G or C (S) while the reference and all Colombian sequences displayed T at that position. However, this mismatch was expected not to affect seriously the overall stability of its hybridization. There was an important finding in the sense primer RdRp_SARSr_F1 which displayed a mismatch at position 15,451 when compared to the hCoV/Colombia/Valle_del_Cauca/79943/2020 genome. This mismatch located at the second base of the 3’ end of the primer is expected to severely affect the primer hybridization and subsequent DNA polymerase-mediated extension. In the present study, we propose to include a degenerate site (R) as follows (RdRP_SARSr-F2 Mod 5’-GTGAAATGGTCATGTGTGGC**R**G-3’). Thus, covering the genetic variability and improving the thermodynamic stability with this genetic variant to avoid false-negative results (Table S2). The nucleotide identity of the Colombian genomes with respect to the Corman Berlin (2020) E Sarbeco and N Sarbeco designs was 100%. These results supported the PAHO recommendation of diagnostic confirmation based on the single E gene in the context of sustained transmission at high levels in this region where other coronavirus species of the subgenus *Sarbecovirus* are expected to be absent (PAHO, 2020).

Another critical characteristic of the primers/probes of the current in-house protocols listed by WHO and assessed in the present study was the codon position at the 3’ end of the sense and antisense primers in coding regions. The major and minor susceptibility to nucleotide substitutions for the third and second codon positions, respectively, is widely known. As any change at the 3’ end can affect the primer hybridization, it is highly recommended that this position aligns with the second (preferable) or first codon position. Some primers and indeed primer sets were found to be coincident with the third codon position. Despite not finding substitutions at these sites in the Colombian strains, that position is expected to be unstable through time.

Genomic data available from this study allowed the *in silico* evaluation/refinement of the protocols for molecular detection of SARS-CoV-2 circulating in Colombia. It is highly recommended to establish routine molecular surveillance of the virus in order to determine the real impact of every mutation in the diagnostic test’s performance (PAHO, 2020).

Finally, the implementation of molecular tests at country-level should be supported by the estimation of the analytical sensitivity (Limit of detection [LOD] in copies/reaction), specificity (Victor M. Corman et al., 2020), and the accumulated genetic variability should be tested during the implementation of a molecular detection protocol including the clinical sensitivity in different biofluids (V. M. Corman et al., 2016), reproducibility, repeatability, inter-operator, inter-instrument, inter-site and inter-batch assays (Hu et al., 2019).

## CONCLUSIONS

Detection of SARS-CoV-2 viral RNA using nucleic acid amplification techniques such as rRT-PCR continues to be the gold standard for the diagnosis of COVID-19 (WHO, 2020d). However, all sequence-based methods are susceptible to nucleotide substitution affecting the oligonucleotide hybridization efficiency and resulting in false negatives. Some of the in-house protocols analyzed in the present study require an experimental evaluation of their performance in the context of virus genetic variability. The genomic data of this study allow the refinement or even the design of more precise and efficient protocols for the molecular detection of the genetic variants of SARS-CoV-2 circulating in Colombia. However, more NGS data from Colombian SARS-CoV-2 will be determinant to a better comprehension of the impact of genetic variability on specific molecular assays of routine use as the virus evolves.

## Data Availability

The data that support the findings of this study are available from the corresponding author, upon reasonable request

## ACKNOWLEDGMENTS

For the joint work with the professionals involved in facing the SARS-CoV-2 pandemic from different aspects and to the Directorates of Public Health Research, Public Health Networks and Surveillance of the National Institute of Health.

## Supplementary material

**Figure S1. Alignment of Pasteur 2020 (France)nCoV IP2 primer sets with Colombian SARS-CoV-2 genomes.**. Genomic positions were estimated according to the SARS-CoV-2 reference genome available at GenBank (NC_045512.2).

**Figure S2. Alignment of Hong Kong (2020) HKU-ORFlb-nspl primer sets with Colombian SARS-CoV-2 genomes**. Genomic positions were estimated according to the SARS-CoV-2 reference genome available at GenBank (NC_045512.2).

**Figure S3. Alignment of Corman Berlin(2020) E Sarbeco primer sets with Colombian SARS-CoV-2 genomes**. Genomic positions were estimated according to the SARS-CoV-2 reference genome available at GenBank (NC_045512.2).

**Figure S4. Alignment of CDC 2020(USA) 2019-nCoV N1 (Fig. S4) and DMC-MH 2020(Thailand)WH-NIC N primer sets with Colombian SARS-CoV-2 genomes**. Genomic positions were estimated according to the SARS-CoV-2 reference genome available at GenBank (NC_045512.2).

**Figure S5. Alignment of CDC 2020(USA) 2019-nCoV N3 and Corman Berlin(2020) N Sarbeco primer sets with Colombian SARS-CoV-2 genomes**. Genomic positions were estimated according to the SARS-CoV-2 reference genome available at GenBank (NC_045512.2).

**Figure S6. Alignment of Nao 2020(Japan)NIID 2019-nCOV N and CDC 2020(USA) 2019-nCoV N2 primer sets with Colombian SARS-CoV-2 genomes**. Genomic positions were estimated according to the SARS-CoV-2 reference genome available at GenBank (NC_045512.2).

**Table S1. Thermodynamic characteristics of the primers/probes showing mismatches with the reference or Colombian SARS-COV-2 sequences**.

**Table S2. Thermodynamic characteristics of RdRP_SARSr-F2 Mod**.

